# Dosing Of Thromboprophylaxis And Mortality In Critically Ill Covid-19 Patients

**DOI:** 10.1101/2020.09.17.20195867

**Authors:** Sandra Jonmarker, Jacob Hollenberg, Martin Dahlberg, Otto Stackelberg, Jacob Litorell, Åsa H Everhov, Hans Järnbert-Pettersson, Mårten Söderberg, Jonathan Grip, Anna Schandl, Mattias Günther, Maria Cronhjort

## Abstract

**Background:** A substantial proportion of critically ill COVID-19 patients develop thromboembolic complications, but it is unclear whether higher doses of thromboprophylaxis are associated with lower mortality rates. The purpose of the study was to evaluate the association of initial dosing strategy of thromboprophylaxis in critically ill COVID-19 patients and the risk of death, thromboembolism, and bleeding.

**Method:** All critically ill COVID-19 patients admitted to two intensive care units in March and April 2020 were eligible. Patients were categorized into three groups according to initial daily dose of thromboprophylaxis; low (2500-4500 IU tinzaparin or 2500-5000 IU dalteparin), medium (>4500 IU but <175 IU/kilogram, kg, of body weight tinzaparin or >5000 IU but <200 IU/kg of body weight dalteparin), and high dose (≥ 175 IU/kg of body weight tinzaparin or ≥200 IU/kg of body weight dalteparin). Thromboprophylaxis dosage was based on local standardized recommendations, not on degree of critical illness or risk of thrombosis. Cox proportional hazards regression was used to estimate hazard ratios with corresponding 95% confidence intervals of death within 28 days from ICU admission. Multivariable models were adjusted for sex, age, body-mass index, Simplified Acute Physiology Score III, invasive respiratory support, and initial dosing strategy of thromboprophylaxis.

**Results:** A total of 152 patients were included; 67 received low, 48 medium, and 37 high dose thromboprophylaxis. Baseline characteristics did not differ between groups. Mortality was lower in high (13.5%) vs medium (25.0%) and low dose thromboprophylaxis (38.8%) groups, p=0.02. The hazard ratio of death was 0.33 (95% confidence intervals 0.13 – 0.87) among those who received high dose, respectively 0.88 (95% confidence intervals 0.43 – 1.83) among those who received medium dose, as compared with those who received low dose thromboprophylaxis. There were fewer thromboembolic events in the high (2.7%) vs medium (18.8%) and low dose thromboprophylaxis (17.9%) groups, p=0.04, but no difference in the proportion of bleeding events, p=0.16.

**Conclusions:** Among critically ill COVID-19 patients with respiratory failure, high dose thromboprophylaxis was associated with a lower risk of death and a lower cumulative incidence of thromboembolic events compared with lower doses.

**Trial registration:** Clinicaltrials.gov NCT04412304 June 2 2020, retrospectively registered

## BACKGROUND

The inflammatory response to Coronavirus disease 19 (COVID-19) seems to trigger thrombotic activation in the venous and the arterial circulation (1, 2). Autopsy findings suggest that this coagulopathy occurs in micro- as well as macrovascular beds (3-5). As many as 17 - 69% of COVID-19 patients in intensive care units (ICU)s suffer from thrombotic events (6-9). This is significantly more than in non-COVID-19 acute respiratory distress syndrome patients (8). For COVID-19 patients, laboratory findings indicate a hypercoagulable state in combination of low grade disseminated intravascular coagulation and thrombotic angiopathy (10, 11) which differ from what is typically seen in sepsis (12). In previous studies, high levels of Fibrin-D-dimer and C-reactive protein (CRP) have been associated with poor outcome in COVID-19 (8, 11-14). In an observational study of hospitalized COVID-19 patients, anticoagulation was associated with improved outcome among mechanically ventilated COVID-19 patients (15), but the optimal choice of dose is yet to be determined. The risk for bleeding with full dose anticoagulants has been described in a small retrospective study where 21% had hemorrhagic events despite anti-factor Xa activity within the therapeutic range for all patients except one (16). At Södersjukhuset, Stockholm, Sweden, the first critically ill COVID-19 patients admitted to the ICUs were treated with low dose thromboprophylaxis. Within a few weeks, after preliminary reports suggesting that a high proportion of COVID-19 patients suffered from thromboembolic events, it was decided to increase the dose of thromboprophylaxis.

The objective of this study was to evaluate the association between dosing strategy of thromboprophylaxis in critically ill COVID-19 patients with respiratory failure and the risk of death, thromboembolism and bleeding.

## METHODS

### Trial overview and patients

In this observational cohort study, all critically ill COVID-19 patients (verified with polymerase chain reaction-positive Severe Acute Respiratory Syndrome Corona Virus 2, SARS-CoV-2) with respiratory failure, admitted to two ICUs in March and April, 2020, at Södersjukhuset, Stockholm, Sweden were eligible for inclusion.

Patients were excluded if discharged the same day as ICU admission, if they had ongoing anticoagulant (AC) therapy prior to ICU due to deep venous thrombosis, DVT, and/or pulmonary embolism, PE, or if they had no initial treatment with thromboprophylaxis in the ICU. Patients with chronic AC therapy at hospital admission, for other reasons than DVT and/or PE, were included in the study.

Data on patients’ demography, comorbidities (International classifications of diseases,10^th^ revision), duration of symptoms, chronic AC therapy, invasive respiratory support, and laboratory values were retrieved from patients’ medical records. Data was automatically and manually extracted by medical doctors and all charts and events were validated by at least one additional medical doctor.

### Dosing strategies of thromboprophylaxis

Patients were categorized into three groups according to initial treatment doses of subcutaneous low-molecular-weight heparin at admission to the ICU. Daily doses of tinzaparin and dalteparin were defined as low dose thromboprophylaxis (2500-4500 international units, IU, tinzaparin or 2500-5000 IU dalteparin), medium dose thromboprophylaxis (>4500 IU but <175 IU/kg of body weight tinzaparin or >5000 IU but <200 IU/kg of body weight dalteparin), and high dose thromboprophylaxis(≥ 175 IU/kg of body weight tinzaparin or ≥200 IU/kg of body weight dalteparin). Patients who received an adjusted dose due to reduced kidney function were classified according to intended dose range.

The choice of dosing strategy followed the local recommendations and were modified over time: In March, low dose thromboprophylaxis was recommended for all COVID-19 patients at both ICUs. In April, the recommendations were altered to medium and then high dose thromboprophylaxis and this was continued throughout the study period in one ICU. In the other ICU, full dose was only used for one week, and then recommendations were altered to medium dose thromboprophylaxis again. All changes in dose were registered with new dose and date.

### Outcomes

The primary outcome was 28-day mortality. Days alive and out of ICU at day 28, the cumulative proportion of thromboembolic and bleeding events within 28 days of ICU admission, and maximum levels of Fibrin-D-dimer were used as secondary outcome measures. Thromboembolic events were PE (verified by computed tomography or by clinical suspicion of PE as cause of deterioration combined with findings of acute strain of the right heart on echocardiography), DVT (verified by ultrasound), ischemic stroke (verified by computed tomography), and peripheral arterial embolism (clinical findings of acute peripheral ischemia). Bleeding events were categorized according to the World health organization (WHO) bleeding scale(17-19): 1) Petechiae, tissue hematoma, oropharyngeal bleeding, 2) Mild blood loss, hematemesis, macroscopic hematuria, hemoptysis, joint bleeding, 3) Gross blood loss requiring red blood cell transfusion and/or hemodynamic instability, 4) Debilitating blood loss, severe hemodynamic instability, fatal bleeding, or central nervous system bleeding.

Baseline laboratory values were obtained from 6 hours before to one hour after ICU admission.

## Statistical analysis

Continuous values for baseline and follow-up data are presented in medians with interquartile range (IQR), while categorical or binary data are shown as numbers and proportions. Differences over categories of the exposure were tested with Kruskal-Wallis for continuous data, and Fisher’s exact for categorical data. In the survival analyses, participants could accrue follow-up time from date of ICU-admission, to date of death, or when 28 days had passed since admission, whichever occurred first. In analyses of thromboembolic and bleeding events, the date of that event also led to censoring of follow-up time. Kaplan-Meier curves were used to estimate the cumulative risk of death, thromboembolic event, and bleeding event, and the log-rank test was used to compare the initial dosing strategies. Cox proportional hazards regression was used to estimate hazard ratios (HR) with corresponding 95% confidence intervals (CI) of death within 28 days from ICU admission. Multivariable models were adjusted for sex, age, body-mass index (BMI), Simplified Acute Physiology Score III (SAPS III), invasive respiratory support (yes/no), and initial dosing strategy of thromboprophylaxis (low, medium and high dose thromboprophylaxis) (20, 21). To assess evidence of non-linearity, the second spline transformation equal to zero was tested as the quantitative covariates were modeled with restricted cubic splines at three knots at fixed percentiles (10^th^, 50^th^ and 90^th^) of the distribution of that covariate (22). As there was no such evidence for age (*P*=0.26), or SAPS III (*P*=0.71), those variables were adjusted for in a continuous fashion. Although no formal evidence, there was an indication of non-linearity between levels of BMI and 28-day mortality (*P*=0.08), why BMI was categorized as </≥ 30 kg/m^2^ with a separate category for missing values (n=6). BMI was flexibly adjusted with restricted cubic splines while missing values were accounted for using chained iterations of multiple imputed data sets (n=20) (23). Although no formal evidence, there was an indication of violation of the assumption of proportional hazards when scaled Schoenfeld residuals were regressed against survival time (*P*=0.06 for high dose thromboprophylaxis). Thus, the time-varying effect was fitted by splitting the follow-up time at 7 days from ICU-admission, and fitting the time varying covariate as an interaction-term with the main exposure. Statistical significance of interaction was tested using the Wald test. To account for other possible temporal changes in treatment over inclusion time, sensitivity analysis was performed with additional adjustment for admission date to ICU. Two-sided P<0.05 was considered statistically significant. Analysis was performed using STATA 13.1 (StataCorp), and R v 3.5.1 (R Core Team (2017). R: A language and environment for statistical computing. R Foundation for Statistical Computing, Vienna, Austria.).

## RESULTS

Out of 165 critically ill COVID-19 patients treated in the ICU due to respiratory failure, 152 remained after exclusion of those with short ICU length of stay (n = 5), ongoing AC therapy at ICU admission due to DVT and/or PE (n = 4), or no initial thromboprophylaxis in the ICU (n = 4) (Figure 1). The reason for not giving thromboprophylaxis to four patients was urgent surgery for one and for the other three their prescriptions were likely overlooked; no reason could be found in the medical record. Patient characteristics are described in Table

**Figure 1.**
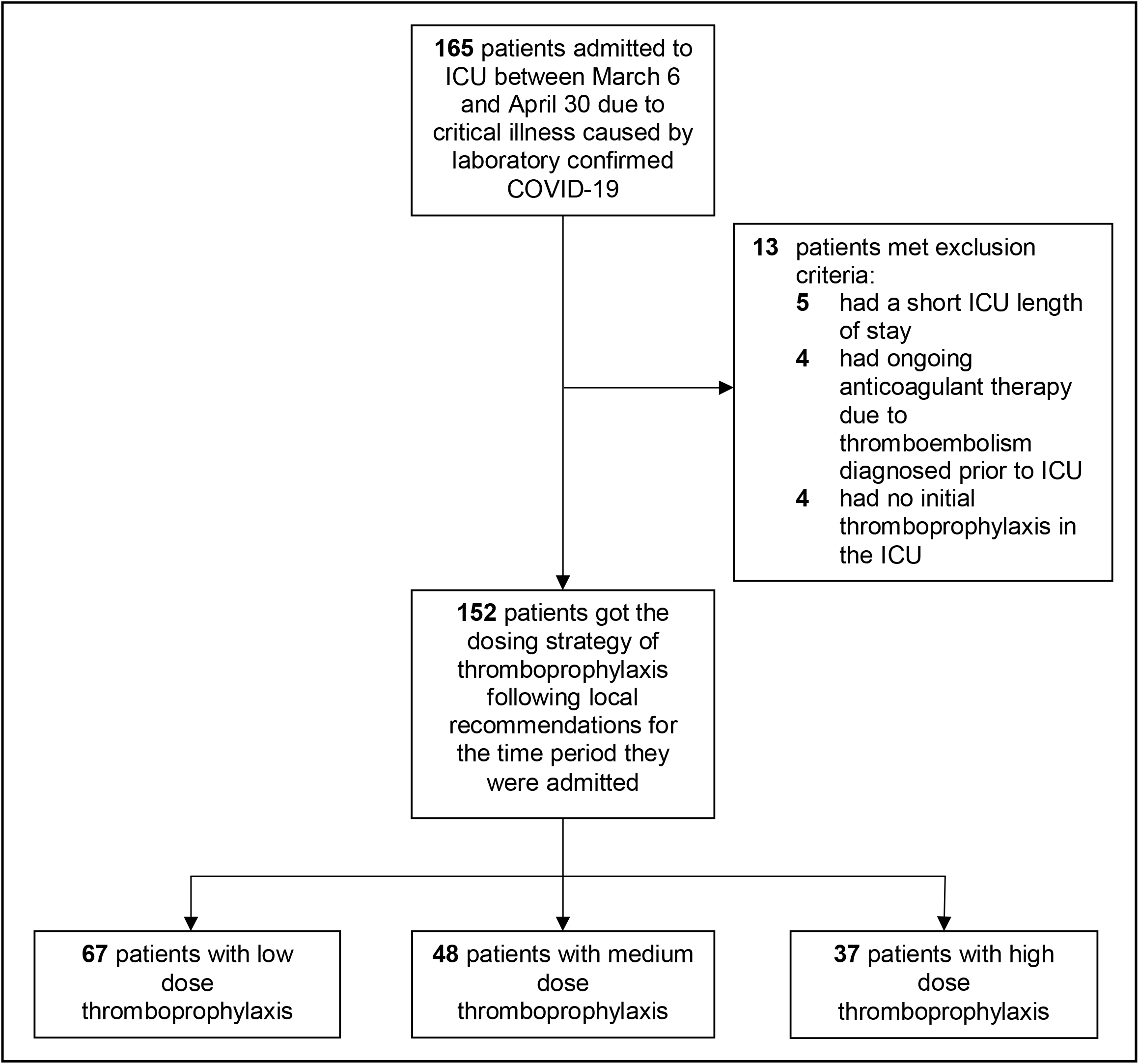
Flow chart. The patients were grouped according to initial dosing strategy of thromboprophylaxis; low (2500-4500 IU of tinzaparin daily, or 2500-5000 IU of dalteparin daily), medium (>4500 IU up to 175 IU/kg of body weight of tinzaparin daily, or >5000 IU up to 200 IU/kg of body weight of dalteparin daily) and high dose thromboprophylaxis (≥175 IU/kg of body weight of tinzaparin daily, or ≥200 IU/kg of body weight of dalteparin daily).

1. The majority of patients included were men (82.2%), the median age was 61 years (IQR 52-69) and the median BMI was 28.4 (IQR 25.8-32.5). The most common comorbidities were hypertension, obstructive lung disease, and diabetes mellitus. Median SAPS III-score was 56 (IQR 50-60) and 68.4% needed invasive respiratory support. The three groups consisted of 67 patients who received low dose thromboprophylaxis, 48 patients who received medium dose thromboprophylaxis and 37 patients who received high dose thromboprophylaxis. Demographic and clinical characteristics did not differ between the three groups. All patients were followed-up until death or until the 28th day after ICU-admission.

### Primary outcome

The 28-day mortality was 38.8% in the group with low dose thromboprophylaxis (26/67), 25.0% with medium (12/48) and 13.5% with high (5/37), (Table 2). The HR of death was 0.33 (95% CI 0.13 – 0.87) among those who received high dose, respectively 0.88 (95% CI, 0.43 – 1.83) among those who received medium dose, as compared with those who received low dose thromboprophylaxis, (Table 3). The cumulative proportion of deaths within the first 28-days from ICU-admission differed between the three groups (P log-rank=0.02). The risk of death did not differ between groups of exposure until 7 days after admission, after which the proportion of deaths increased in the low dose thromboprophylaxis group compared to the other groups (Figure 2A). When follow-up time was split and the time-varying covariate was fitted as an interaction term with the exposure (Table 4), the HR of high dose thromboprophylaxis was 0.08 (95% CI, 0.01 – 0.62) day 7 to 28, compared with low dose thromboprophylaxis the same time period, while no differences were observed during day 0 to 7. The interaction term itself was not significant overall (*P*=0.18). Among the first half of admissions (before April 11, 2020), 74.7% (n=59) received low dose and 2.5% (n=2) high dose thromboprophylaxis. Corresponding proportions in the second half of admissions was 11.0% (n=8), respectively 48% (n=35). In analysis with additional adjustment for median admission date, the HR of death was 0.51 (95% CI, 0.14 – 1.89) of high vs low dose thromboprophylaxis, respectively 0.63 (95% CI, 0.24 – 1.64) comparing the second to the first half of admissions.

**Table 1.** Baseline characteristics by initial dosing strategy of thromboprophylaxis. Baseline characteristics of 152 patients admitted to the intensive care unit due to COVID-19 at Södersjukhuset, Stockholm, March 6 to April 30, 2020, by initial dosing strategy with tinzaparin/dalteparin as thromboprophylaxis.

|  | Total<br>(n=152) | Initial dosing strategy of thromboprophylaxis |  |  | P-value<sup>d</sup> |
| --- | --- | --- | --- | --- | --- |
|  |  | Low dose<sup>a</sup><br>(n=67) | Medium dose<sup>b</sup><br>(n=48) | High dose<sup>c</sup><br>(n=37) |  |
| Age, years | 61 (52 – 69) | 63 (52 – 71) | 58 (51 – 66) | 63 (54 – 70) | 0.39 |
| Male sex, No.(%) | 125 (82.2) | 59 (88.1) | 35 (72.9) | 31 (83.8) | 0.12 |
| BMI, kg/m<sup>2</sup> | 28.4 (25.8 – 32.5) | 27.7 (25.5 – 30.6) | 29.4 (26.5 – 34.3) | 28.4 (25.1 – 32.8) | 0.10 |
| BMI ≥30 kg/m<sup>2</sup>, No.(%) | 58 (39.7) | 20 (31.8) | 23 (48.9) | 15 (41.7) | 0.18 |
| Tobacco use |  |  |  |  |  |
| Never, No.(%) | 79 (52.0) | 32 (47.8) | 28 (58.3) | 19 (51.4) | 0.53 |
| Former smoker, No.(%) | 49 (32.2) | 24 (35.8) | 12 (25.0) | 13 (35.1) | 0.40 |
| Current smoker, No.(%) | 7 (4.6) | 3 (4.5) | 3 (6.3) | 1 (2.7) | 0.79 |
| Comorbidities |  |  |  |  |  |
| Hypertension, No.(%) | 69 (45.4) | 32 (47.8) | 20 (41.7) | 17 (46.0) | 0.83 |
| Diabetes without complications, No.(%) | 19 (12.5) | 8 (11.9) | 6 (12.5) | 5 (13.5) | 0.99 |
| Diabetes with complications, No.(%) | 6 (4.0) | 4 (6.0) | 0 (0.0) | 2 (5.4) | 0.24 |
| Obstructive pulmonary disease, No.(%) | 30 (19.7) | 12 (17.9) | 11 (22.9) | 7 (18.9) | 0.81 |
| Ischemic heart disease, No.(%) | 12 (7.9) | 6 (9.0) | 2 (4.2) | 4 (10.8) | 0.51 |
| Renal failure, No.(%) | 9 (5.9) | 6 (9.0) | 3 (6.3) | 0 (0.0) | 0.17 |
| Liver disease, No.(%) | 3 (2.0) | 1 (1.5) | 1 (2.1) | 1 (2.7) | 0.99 |
| Immunosuppression, No.(%) | 8 (5.3) | 5 (7.5) | 1 (2.1) | 2 (5.4) | 0.53 |
| Malignancy, No.(%) | 9 (5.9) | 6 (9.0) | 1 (2.1) | 2 (5.4) | 0.34 |
| Psychiatric disorder, No.(%) | 26 (17.1) | 11 (16.4) | 9 (18.8) | 6 (16.2) | 0.96 |
| Chronic medication |  |  |  |  |  |
| Vitamin K antagonist, No.(%) | 1 (0.7) | 0 (0.0) | 0 (0.0) | 1 (2.7) | 0.24 |
| Direct oral anticoagulation, No.(%) | 8 (5.3) | 3 (4.5) | 1 (2.1) | 4 (10.8) | 0.24 |
| Antiplatelet therapy, No.(%) | 15 (9.9) | 9 (13.4) | 3 (6.3) | 3 (8.1) | 0.47 |
| Duration of symptoms prior to admission |  |  |  |  |  |
| Hospital admission, days | 9 (7 – 12) | 9 (6 – 13) | 8 (6 – 12) | 10 (7 – 14) | 0.18 |
| ICU admission, days | 12 (8 – 14) | 12 (8 – 14) | 10 (8 – 14) | 13 (10 – 14) | 0.13 |
| SAPS III-score | 56 (50 – 60) | 57 (53 – 64) | 54 (49 – 59) | 55 (53 – 60) | 0.25 |
| <50, No.(%) | 34 (22.4) | 15 (22.4) | 13 (27.1) | 6 (16.2) | 0.53 |
| 50-59, No.(%) | 75 (44.7) | 31 (46.3) | 25 (52.1) | 19 (51.4) | 0.76 |
| ≥60, No.(%) | 43 (28.3) | 21 (31.3) | 10 (20.8) | 12 (32.4) | 0.41 |
| Invasive Respiratory support, No.(%) | 104 (68.4) | 50 (74.6) | 34 (70.8) | 20 (54.1) | 0.09 |
| Coagulation, Inflammatory and creatinine tests at baseline |  |  |  |  |  |
| Fibrin-D-dimer, mg/L FEU | 1.5 (0.90 – 3.9) | 1.8 (1.1 – 8.4) | 1.1 (0.7 – 2.4) | 1.3 (0.7 – 2.9) | 0.12 |
| CRP, mg/L | 195 (134 – 270) | 198 (134 – 268) | 203 (151 – 255) | 183 (130 – 287) | 0.81 |
| Hemoglobin, g/L | 129 (118 – 136) | 127 (118 – 137) | 130 (119 – 137) | 127 (113 – 135) | 0.39 |
| Creatinine, µmol/L | 70 (57 – 88) | 74 (61 – 92) | 69 (48 – 87) | 62 (50 – 74) | 0.12 |
| Platelet count, 10<sup>9</sup>/L | 264 (195 – 339) | 255 (183 – 332) | 261 (219 – 318) | 302 (225 – 391) | 0.16 |
| INR, | 1.1 (1.0 – 1.1) | 1.1 (1.0 – 1.1) | 1.1 (1.0 – 1.1) | 1.0 (1.0 – 1.2] | 0.89 |
| APTT, seconds | 26 (24 – 29) | 25 (24 – 29) | 26 (23 – 27) | 27 (25 – 30) | 0.32 |
| Fibrinogen, g/L | 6.7 (5.5 – 8.0) | 6.7 (5.6 – 7.7) | 7.2 (6.4 – 8.3) | 6.7 (5.4 – 7.7) | 0.47 |

Values are medians (interquartile range) unless otherwise indicated. APTt indicates activated partial
thromboplastin time; BMI, body mass index; FEU, Fibrinogen equipotent unit; CRP, C-reactive protein; ICU,
intensive care unit; INR, Prothrombin Time International Normalized Ratio; SAPS III, Simplified Acute
Physiology Score III.
<sup>a</sup> tinzaparin, 2500-4500 IU daily; or dalteparin, 2500-5000 IU daily
<sup>b</sup> tinzaparin, >4500 IU daily to <175 IU/kg of body weight daily; or dalteparin, >5000 IU daily to <200 IU/kg of body weight daily
<sup>c</sup> tinzaparin, ≥175 IU/kg of body weight daily; or dalteparin, ≥200 IU/kg of body weight daily
<sup>d</sup> P-values for differences across exposure categories were obtained using Fisher's exact test for categorical and Kruskal-Wallis test for continuous data.

**Table 2.**
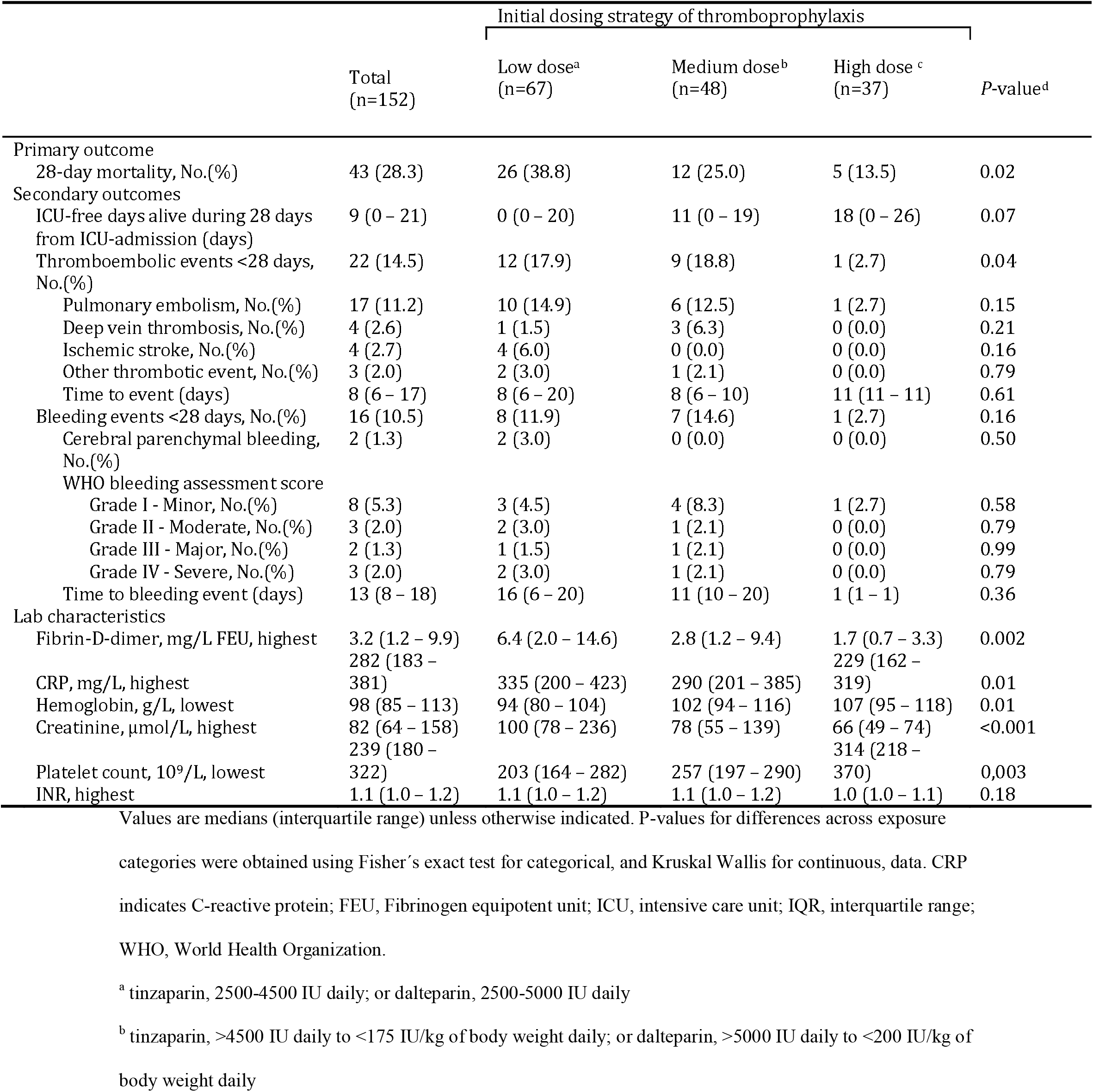
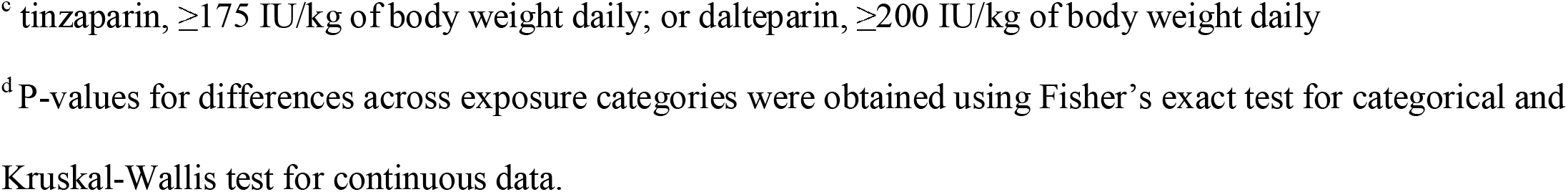
Outcomes by initial dosing strategy of thromboprophylaxis. Primary and secondary outcomes during the first 28 days among 152 patients admitted to the intensive care unit due to COVID-19 at Södersjukhuset, Stockholm, March 6 to April 30, 2020, by initial dosing strategy with tinzaparin/dalteparin as thromboprophylaxis.

**Table 3.** Risk of death by initial dosing strategy of thromboprophylaxis. Risk of death during the first 28 days among 152 patients admitted to the intensive care unit due to COVID-19 at Södersjukhuset, Stockholm, March 6 to April 30, 2020, by initial dosing strategy with tinzaparin/dalteparin as thromboprophylaxis.

| Initial dosing strategy of thromboprophylaxis | No. of patients | Events / person-days | IR per 1.000 person-days (95% CI) | HR (95% CI) of death ≤28 days |  |  |
| --- | --- | --- | --- | --- | --- | --- |
|  |  |  |  | Univariable model | Multivariable model<sup>a</sup> | Multivariable imputed model<sup>b</sup> |
| High dose<sup>c</sup> | 37 | 5 / 923 | 5.4 (2.3 – 13.0) | 0.31 (0.12 – 0.82) | 0.33 (0.13 – 0.87) | 0.30 (0.11 – 0.81) |
| Medium dose<sup>d</sup> | 48 | 12 / 1182 | 10.2 (5.8 – 17.9) | 0.59 (0.30 – 1.16) | 0.88 (0.43 – 1.83) | 0.87 (0.42 – 1.82) |
| Low dose<sup>e</sup> | 67 | 26 / 1453 | 17.9 (12.2 – 26.3) | 1.00 (Ref.) | 1.00 (Ref.) | 1.00 (Ref.) |

CI indicates Confidence Interval; IR, Incidence Rate; HR, Hazard Ratio.
<sup>a</sup> Adjusted for sex, age (continuously), body-mass index ( $</\geq 30$ kg/m<sup>2</sup> and missing [n=6]), invasive mechanical ventilation (yes/no), and Simplified Acute Physiology Score III (continuously)
<sup>b</sup> Adjusted like the multivariable model but with body-mass index imputed due to missing values (n=6), and flexibly modeled with restricted cubic splines at three knots over the percentile (10<sup>th</sup>, 50<sup>th</sup>, and 90<sup>th</sup>) distribution of body-mass index in the population.
<sup>c</sup> tinzaparin, $\geq 175$ IU/kg of body weight per daily, or dalteparin, $\geq 200$ IU/kg of body weight daily
<sup>d</sup> tinzaparin, $>4500$ IU daily to $<175$ IU/kg of body weight daily, or dalteparin, $>5000$ IU daily to $<200$ IU/kg of body weight daily
<sup>e</sup> tinzaparin, 2500-4500 IU daily, or dalteparin, 2500-5000 IU daily

**Table 4.**
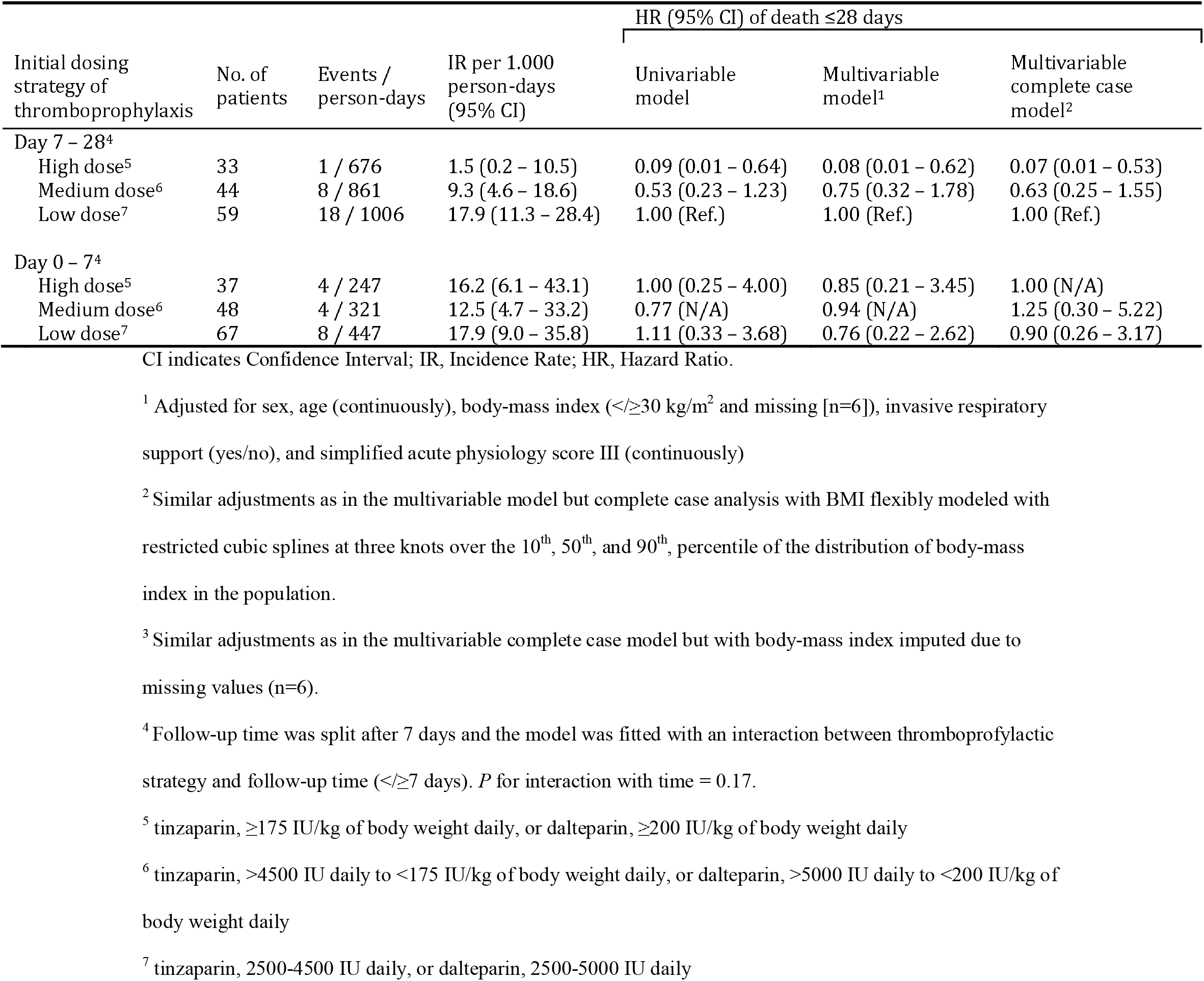
The risk of death when follow-up was split at first week. Risk of death during the first 28 days, with follow-up split at the first week, among 152 patients admitted to the intensive care unit due to COVID-19 at Södersjukhuset, Stockholm, March 6 to April 30, 2020, by initial treatment regime with tinzaparin/dalteparin as thromboprophylaxis.

| Initial dosing strategy of thromboprophylaxis | No. of patients | Events / person-days | IR per 1.000 person-days (95% CI) | HR (95% CI) of death ≤28 days |  |  |
| --- | --- | --- | --- | --- | --- | --- |
|  |  |  |  | Univariable model | Multivariable model<sup>1</sup> | Multivariable complete case model<sup>2</sup> |
| Day 7 – 28<sup>4</sup> |  |  |  |  |  |  |
| High dose<sup>5</sup> | 33 | 1 / 676 | 1.5 (0.2 – 10.5) | 0.09 (0.01 – 0.64) | 0.08 (0.01 – 0.62) | 0.07 (0.01 – 0.53) |
| Medium dose<sup>6</sup> | 44 | 8 / 861 | 9.3 (4.6 – 18.6) | 0.53 (0.23 – 1.23) | 0.75 (0.32 – 1.78) | 0.63 (0.25 – 1.55) |
| Low dose<sup>7</sup> | 59 | 18 / 1006 | 17.9 (11.3 – 28.4) | 1.00 (Ref.) | 1.00 (Ref.) | 1.00 (Ref.) |
| Day 0 – 7<sup>4</sup> |  |  |  |  |  |  |
| High dose<sup>5</sup> | 37 | 4 / 247 | 16.2 (6.1 – 43.1) | 1.00 (0.25 – 4.00) | 0.85 (0.21 – 3.45) | 1.00 (N/A) |
| Medium dose<sup>6</sup> | 48 | 4 / 321 | 12.5 (4.7 – 33.2) | 0.77 (N/A) | 0.94 (N/A) | 1.25 (0.30 – 5.22) |
| Low dose<sup>7</sup> | 67 | 8 / 447 | 17.9 (9.0 – 35.8) | 1.11 (0.33 – 3.68) | 0.76 (0.22 – 2.62) | 0.90 (0.26 – 3.17) |

CI indicates Confidence Interval; IR, Incidence Rate; HR, Hazard Ratio.
<sup>1</sup> Adjusted for sex, age (continuously), body-mass index ( $</\geq 30$ kg/m<sup>2</sup> and missing [n=6]), invasive respiratory support (yes/no), and simplified acute physiology score III (continuously)
<sup>2</sup> Similar adjustments as in the multivariable model but complete case analysis with BMI flexibly modeled with restricted cubic splines at three knots over the 10<sup>th</sup>, 50<sup>th</sup>, and 90<sup>th</sup>, percentile of the distribution of body-mass index in the population.
<sup>3</sup> Similar adjustments as in the multivariable complete case model but with body-mass index imputed due to missing values (n=6).
<sup>4</sup> Follow-up time was split after 7 days and the model was fitted with an interaction between thromboprophylactic strategy and follow-up time ( $</\geq 7$ days). *P* for interaction with time = 0.17.
<sup>5</sup> tinzaparin, $\geq 175$ IU/kg of body weight daily, or dalteparin, $\geq 200$ IU/kg of body weight daily
<sup>6</sup> tinzaparin, $>4500$ IU daily to $<175$ IU/kg of body weight daily, or dalteparin, $>5000$ IU daily to $<200$ IU/kg of body weight daily
<sup>7</sup> tinzaparin, 2500-4500 IU daily, or dalteparin, 2500-5000 IU daily

**Figure 2.**
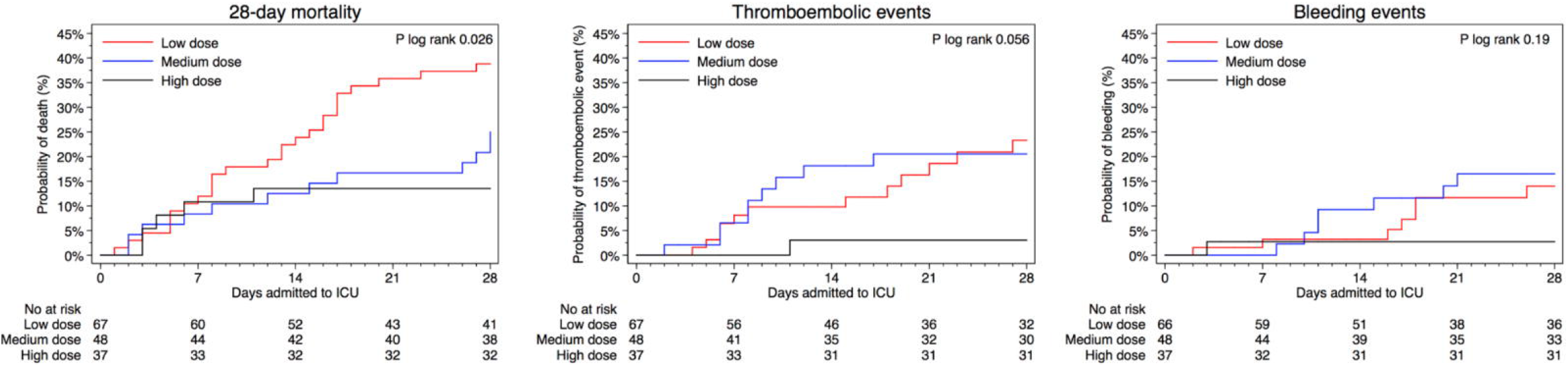
Kaplan-Meier plots of outcomes by initial dosing strategy of thromboprophylaxis. Kaplan-Meier plot of (a) 28-day survival, (b) thromboembolic events, and (c) bleeding events, among 152 patients admitted to the ICU due to COVID-19 between March 6 and April 30, 2020. By thromboprophylactic anticoagulant strategy with tinzaparin/dalteparin: The red line represent low dose thromboprophylaxis (2500-4500 IU of tinzaparin daily, or 2500-5000 IU of dalteparin daily), the blue line represent medium dose thromboprophylaxis (>4500 IU to <175 IU/kg of body weight of tinzaparin daily, or >5000 IU to <200 IU/kg of body weight of dalteparin daily), and the black line represent high dose thromboprophylaxis (≥175 IU/kg of body weight of tinzaparin daily, or ≥200 IU/kg of body weight of dalteparin daily). Thromboembolic events in (B) are defined as pulmonary embolism, deep vein thrombosis, ischemic stroke, or peripheral arterial embolism. Hemorrhagic events in (c) are defined as grade 1-4 in the WHO bleeding scale

### Secondary outcomes

The median number of ICU free days alive during the first 28 days were 0 (IQR 0-22) with low, 11 (IQR 0-19) with medium and 18 (IQR 0-26) with high dose thromboprophylaxis, (p=0.07). The proportion of thromboembolic events for the three groups was 17.9% with low, 18.8% with medium and 2.7% with high dose thromboprophylaxis, (p= 0.04). The proportion of bleeding events was 11.9% with low, 14.6% with medium and 2.7% with high dose thromboprophylaxis (p=0.16). Cumulative proportions of thromboembolic events, and bleeding events, are depicted in figure 2B, and figure 2C, respectively. In the low dose thromboprophylaxis group, four patients suffered ischemic stroke, and two had minor intracranial hemorrhage. There were five major or severe bleeding events, three with low and two with medium dose thromboprophylaxis (Table 2).

### Changes in dose

Dosing of thromboprophylaxis was registered and followed over time as seen in figure 3. Of the 152 patients 69 (45.4%) had one or more dose changes during the ICU stay and for 64 patients (42.1%) the changes included an increase. There were 9 (13.0%) patients who had dose adjustments including both increases and reductions and for 5 (3.3%) patients the adjustment was a reduction compared to initial dose. The median treatment time before a change in dose was 4 (2-7) days. Median duration of thromboprophylaxis was 8 (3-17) days.

**Figure 3.**
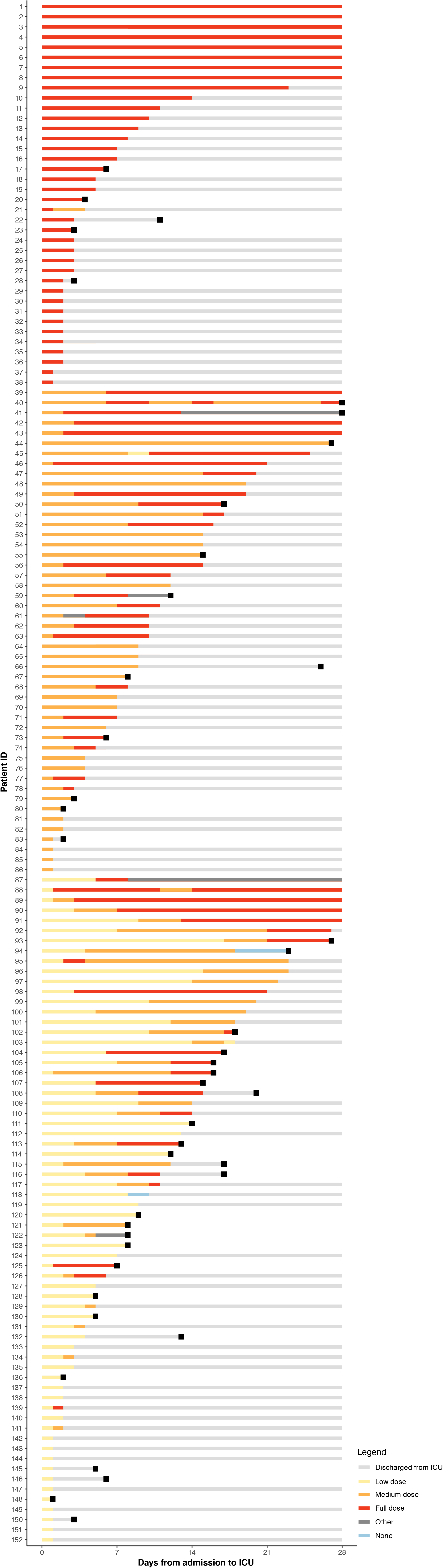
Dosing of thromboprophylaxis. Dosing of thromboprophylaxis for all 152 patients from admission to intensive care until discharge, death or a maximum of 28 days. Each line represent one patient and changes in dosing are represented by altered color. Yellow corresponds to low dose thromboprophylaxis (2500-4500 IU of tinzaparin daily, or 2500-5000 IU of dalteparin daily), orange corresponds to medium dose thromboprophylaxis (>4500 IU to <175 IU/kg of body weight of tinzaparin daily, or >5000 IU to <200 IU/kg of body weight of dalteparin daily), and red corresponds to high dose thromboprophylaxis (≥175 IU/kg of body weight of tinzaparin daily, or ≥200 IU/kg of body weight of dalteparin daily). Grey corresponds to other anticoagulant therapies (direct oral anticoagulation, vitamin K antagonists, heparin infusion). Blue line corresponds to no anticoagulant therapy. Grey line is time after discharge for which no doses were registered. Black squares marks time of death.

### Laboratory results

While there were no differences in Fibrin-D-dimer, CRP, hemoglobin (Hb), platelet count and creatinine values at baseline (Table 1), the maximum levels during the first 28 days differed (Table 2). The highest maximum level of Fibrin-D-Dimer was in the group receiving low dose thromboprophylaxis (6.4 (2.0 – 14.6), the second highest in the group with medium dose (2.8 (1.2 – 9.4)), and the lowest in the group with high dose thromboprophylaxis (1.7 (0.7 – 3.3)), p = 0.002. The pattern was similar for CRP (p = 0.01) and creatinine (p <0.001). For Hb (p=0.01) and platelet count (p=0.003) the lowest values were found in the group which received low dose thromboprophylaxis and the highest in the group with high dose. Values over time are presented in figure 4.

**Figure 4.**
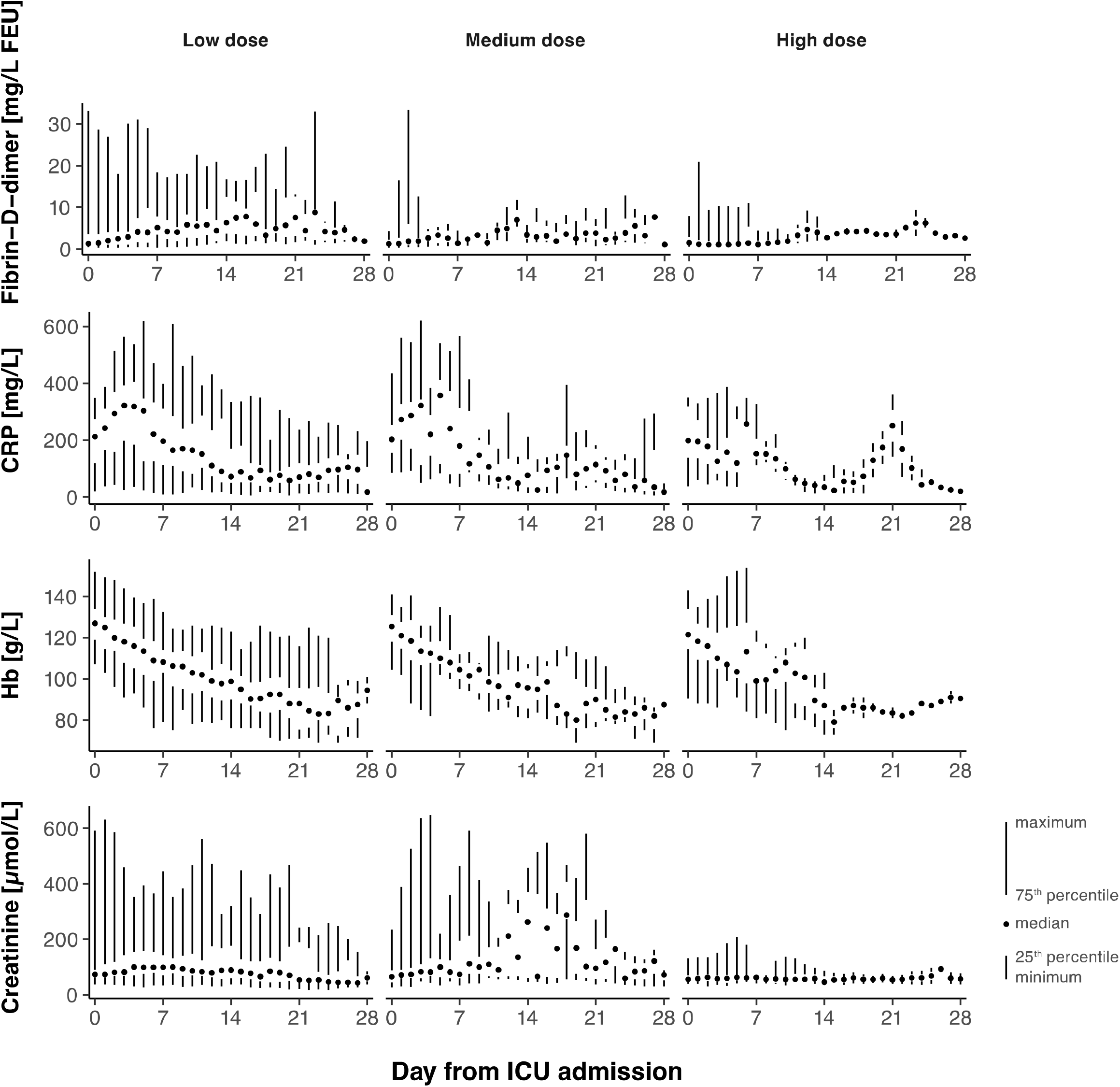
Laboratory markers by initial dosing strategy of thromboprophylaxis. Laboratory markers Fibrin-D-Dimer, C-reactive protein (CRP), hemoglobin concentration, and creatinine as a function of day from admission are shown as median, interquartile range and range. Columns are low dose thromboprophylaxis (2500-4500 IU of tinzaparin daily, or 2500-5000 IU of dalteparin daily), medium dose thromboprophylaxis (>4500 IU to <175 IU/kg of body weight of tinzaparin daily, or >5000 IU to <200 IU/kg of body weight of dalteparin daily), and high dose thromboprophylaxis (≥175 IU/kg of body weight of tinzaparin daily, or ≥200 IU/kg of body weight of dalteparin daily).

## DISCUSSION

To the best of our knowledge, this is the first study that has compared three different doses of anticoagulants to critically ill patients with Covid-19 in relation to 28-day mortality, thromboembolism, and bleeding events. Our main finding is that we estimated a 67% decreased risk of death within the first 28 days among those who received high, as compared with those who received low dose thromboprophylaxis. Also, high dose thromboprophylaxis was associated with a lower incidence of thromboembolic events while no increased risk of bleeding was observed. The 28-day mortality was 38.8% with low, 25.0% with medium and 13.5% with high dose thromboprophylaxis which could indicate a dose-response relationship.

The dose of thromboprophylaxis was based on local standardized recommendations and not on degree of critical illness or risk for thrombosis. This is illustrated by the fact that there were no differences in SAPS III score or Fibrin-D-dimer and CRP levels at baseline, and that patient characteristics were similar between groups. As the local recommendations regarding thromboprophylaxis changed over time 45.4% of patients had a change in dose with the vast majority being increases and only 3.3 % having a reduction from initial dose at some point during the ICU-stay. This suggests that the observed higher mortality in the groups with lower doses could have been even higher if the initial dosing had been sustained.

Our results are in congruence with another study where lower mortality was seen when mechanically ventilated COVID-19 patients were treated with higher doses of anticoagulation (15).

In a retrospective study by Trinh et al., still in pre-print, outcomes for 244 COVID-19 patients on invasive respiratory support were investigated with respect to dose of thromboprophylaxis. This study reports a 79% decreased mortality in therapeutic vs prophylactic anticoagulation, but with a trend towards increased risk of bleeding in the therapeutic group, contrary to our findings of similar proportions of bleeding events in all groups.

It is still unclear how SARS-CoV-2 affects coagulation, and to what degree the hypercoagulable state contributes to the observed respiratory failure, but it is conceivable that early thrombus formation plays an important part in the rapid progression of the pulmonary disease process. Subclinical bleeding could potentially impact the safety of high dose thromboprophylaxis. Hemoglobin values are known to drop during intensive care. As many as 97% of patients are anemic after a week’s ICU care(24, 25). Decreased erythropoiesis, frequent blood sampling and bleeding are known reasons why anemia is common in ICU patients (26). Interestingly, no signs were found of more severe anemia in patients with higher doses of anticoagulation, perhaps because COVID-19 patients are hypercoagulable and might not bleed easily despite high dose thromboprophylaxis. There was even a higher proportion of bleeding in the group with low dose, 11.9%, vs high dose, 2.7% but no significant difference. This result differs from a recent report of 21% incidence of bleeding among 92 critically ill COVID-19 patients, of which 43 had prophylactic dose and 49 had full dose anticoagulation (16).

### Strengths and limitations

The present study lacks the rigor of a prospective randomized design. The study groups thus reflect the progression from low to high dose thromboprophylaxis based on modifications of local clinical guidelines over time. As the study is observational, other factors might have affected outcome. During the study period the ventilation strategy was changed from a classical acute respiratory distress syndrome strategy with low tidal volumes, fluid restriction, and heavy sedation to a more liberal strategy allowing higher tidal volumes, more fluids and less sedation. However, in analysis with additional adjustment for median admission date, the risk of death was still 49% lower (although not statistically significant) among those who received high dose compared with low dose prophylaxis. Furthermore, patients were grouped according to initial dose of thromboprophylaxis at admission to the ICU, and outcomes in relation to total dose thromboprophylaxis received (including changes in dosing strategy during the ICU care) have not been analyzed. There was also a lower proportion of patients on invasive ventilation in the group with high dose, though not statistically significant, and this was adjusted for in the statistical analysis.

Due to the heavy work load during the pandemic, risk of complications during intrahospital transportation of critically ill patients due to respiratory instability, and the risk of viral contamination of radiology suites, it was not always possible to perform computed tomography-scans to diagnose PE. As the number of PE might be underestimated, decision was made to analyze mortality as primary outcome.

The results regarding risk of bleeding should be interpreted with caution due to low numbers, limiting statistical power. Thus, if high dose thromboprophylaxis is used, it is important to continue to monitor the patients closely for potential bleeding.

## CONCLUSIONS

Among critically ill COVID-19 patients with respiratory failure, high dose thromboprophylaxis was associated with a lower risk of death and a lower cumulative incidence of thromboembolic events compared with lower doses. This study suggests that high dose thromboprophylaxis should be tested in a randomized trial in critically ill COVID-19 patients.

## Data Availability

The datasets generated and analyzed during current study are not publicly available due to patient records regulations but can be made available from corresponding author on request.

## DECLARATIONS

### Ethics approval and consent to participate

The study was approved by the regional ethical review board in Uppsala, Sweden, (Dnr: 2020-01302, amendment 2020-02890) and informed consent was waived.

### Consent for publication

Not applicable

### Availability of data and materials

The datasets generated and analyzed during current study are not publicly available due to patient records’ regulations but can be made available from corresponding author on request.

## Author’s Contributions

Concept: S. Jonmarker, J. Hollenberg, M. Dahlberg, O. Stackelberg, J. Litorell, Å. H. Everhov, A. Schandl, M. Cronhjort

Data collection: S. Jonmarker, J. Hollenberg, M. Dahlberg, O. Stackelberg, J. Litorell, Å. H. Everhov, A. Schandl, M. Günther, M. Cronhjort

Data analysis: O. Stackelberg, H. J. Pettersson

Interpretation: S. Jonmarker, J. Hollenberg, M. Dahlberg, O. Stackelberg, J. Litorell, Å. H. Everhov, A. Schandl, M. Söderberg, H. J. Pettersson, M. Cronhjort

Drafting manuscript: S. Jonmarker, M. Cronhjort

Editing manuscript: S. Jonmarker, J. Hollenberg, M. Dahlberg, O. Stackelberg, J. Litorell, Å. H. Everhov, A. Schandl, M. Söderberg, H. J. Pettersson, M. Günther, M. Cronhjort

M. Cronhjort, S. Jonmarker and O. Stackelberg had full access to all the data in the study and take responsibility for the integrity of the data and the accuracy of the data analysis.

## Acknowledgements

We would like to thank W. Müller for his whole-hearted support in data collection from electronic sources.

## LIST OF ABBRIVIATIONS

COVID: coronavirus disease
ICU: intensive care units
CRP: c-reactive peptide
SARS-CoV-2: Severe acute respiratory distress syndrome Coronavirus 2
AC: anticoagulant
DVT: deep vein thrombosis
PE: pulmonary embolism
IU: international units
WHO: world health organization
IQR: interquartile range
HR: hazard ratio
CI: confidence intervals
BMI: body mass index
SAPS: simplified acute physiology score
Hb: hemoglobin
Kg: kilogram
APTt: activated partial thromboplastin time
INR: prothrombin time international normalized ratio
IR: incidence Rate
FEU: Fibrinogen equipotent unit

